# AortoBVI: A Machine Learning-derived Risk Stratification of Ascending Aortic Aneurysms Predictable by Imaging

**DOI:** 10.64898/2026.11.16.26368253

**Authors:** Jennifer C.Y. Chung, Farshad Tajeddini, Daniella Eliathamby, Nitish Bhatt, Hanfei Su, Hijun Seo, David Holt, Maral Ouzounian, Kate Hanneman, Chris McIntosh, Craig A. Simmons

**Affiliations:** Institute of Biomedical Engineering, University of Toronto, Toronto, ON, Canada; Peter Munk Cardiac Centre, University Health Network, University of Toronto, Toronto, ON, Canada; Temerty Faculty of Medicine, University of Toronto, Toronto, ON, Canada; Department of Mechanical & Industrial Engineering, University of Toronto, Toronto, ON, Canada; Department of Medical Imaging, University Medical Imaging Toronto, University of Toronto, Toronto, ON; Translational Biology and Engineering Program, Ted Rogers Centre for Heart Research, Toronto, Canada

## Abstract

**Background:** Current diameter-based risk stratification for ascending aortic aneurysms incompletely captures patient-specific dissection risk. Here, we apply machine learning to ex vivo aortic biomechanics to define a tissue-derived risk continuum, termed aortic biomechanical vulnerability index (AortoBVI), and use magnetic resonance imaging (MRI) to predict this index in vivo.

**Methods:** Ascending aortic tissue from 353 aneurysms, 45 normal aortas, and 46 acute aortic dissections underwent biomechanical testing. Biomechanical features were used to train a classifier distinguishing normal-like, intermediate, and dissection-like biomechanical phenotypes. Magnetic resonance imaging was performed on 38 aneurysm patients pre-operatively and 8 normal controls.

**Results:** The classifier achieved an area under the curve (AUC) of 0.901 (95% CI 0.836-0.954). Among 38 aneurysms with pre-operative MRI, a model combining age, aortic stiffness index (ASI), and aortic pulse wave velocity (aPWV) estimated AortoBVI (R^2^ = 0.580), identifying dissection-like aneurysms with an AUC of 0.928 (95% CI 0.826–0.994. These results significantly outperform aortic diameter (R^2^ = 0.116, AUC 0.620).

**Conclusion:** These findings demonstrate substantial biomechanical heterogeneity at similar aortic diameters and support biomechanics-based risk stratification of ascending aortic aneurysms.

**Clinical Perspective:** *What is New?:*

- AortoBVI integrates five ex vivo biomechanical measurements into a continuous index anchored to normal and acute-dissection tissue phenotypes.
- Among 353 ascending aortic aneurysms, tissue-derived AortoBVI revealed substantial biomechanical heterogeneity despite similar aortic diameters.
- A model combining age with MRI-derived aortic stiffness index and aortic pulse wave velocity estimated tissue-derived AortoBVI more accurately than maximum aortic diameter.

*What Are the Clinical Implications?:*

- Aortic diameter alone does not fully capture the biomechanical variation among ascending aortic aneurysms.
- MRI-based AortoBVI could provide a non-invasive approach to biomechanical phenotyping that complements diameter-based assessment.
- Biomechanically phenotyping patients with ascending aortic aneurysms using AortoBVI may help individualize surveillance intensity and the timing of surgery.

## Introduction

Aneurysms of the ascending aorta affect up to 4% of the general population^1,2^ and place patients at risk of acute type A aortic dissection (ATAAD), with catastrophic consequences including death and organ malperfusion. Replacement of the ascending aorta eliminates this lifetime risk. However, population-based estimates of ATAAD in patients with aortic aneurysm are only 2% per patient-year^3^. Therefore, patient selection for surgery, which carries a similar risk of death and stroke, remains the central conundrum in treating aortic disease.

Current clinical guidelines for elective surgery are based on maximum aneurysm diameter^4,5^, despite its limited ability to predict dissection risk. In the International Registry of Acute Aortic Dissection, 59% of 591 patients with ATAAD had a maximum aortic diameter less than the current intervention threshold of 55 mm and 40% had a diameter <50 mm^6^, with similar findings reported in single center contemporary studies^7,8^. In studies accounting for the acute changes in aortic dimensions with the dissection event, maximum pre-dissection aortic diameter was estimated to be <55 mm in 72-92% of patients^9–11^. Our group and others have shown that diameter provides limited insight into tissue microstructure and biomechanical behavior^12–15^. Other metrics that more directly describe patient-specific aortic wall fragility and individualized risk of dissection and rupture are therefore needed.

Bench-top ex vivo mechanical testing uses several biomechanical parameters to describe the spectrum of material properties of the diseased aortic wall. These include delamination strength, derived by peel testing the aortic wall to simulate dissection^16^, and elastic modulus, which is increased in diseased tissue^17^. Aneurysmal tissue also exhibits reduced performance in returning stored mechanical energy to blood flow (the Windkessel function) each cardiac cycle. Energy loss, describing the hysteresis between loading and unloading of a stress-strain curve, has been found to correlate with aortic wall histopathology and delamination strength^13,18^.

For clinical translation, aortic biomechanical parameters require predictive value for aortic dissection and accurate non-invasive estimation, such as via medical imaging^19^. Several parameters have been suggested to measure vessel stiffness in vivo, including strain^20,21^, strain-based parameters (e.g., distensibility, compliance, and aortic stiffness index (ASI))^22–25^, and aortic pulse wave velocity (aPWV)^26–28^. Unfortunately, none are in clinical use as their correlations with ex vivo biomechanics and clinically relevant endpoints are underexplored, hindering their interpretation and application^19^.

To address these limitations, we aimed to risk-stratify patients into clinically meaningful categories based on their ex vivo aortic biomechanics using machine learning. We also explored whether magnetic resonance imaging (MRI)-derived parameters could reproduce this risk stratification, providing a non-invasive method of personalized risk assessment for patients with aortic aneurysms.

## Methods

### Study Design and Patient population

This prospective cohort study was approved by the research ethics board of the University Health Network, Toronto (16-6285, July 27, 2017). Included for ex vivo mechanical testing of aortic tissue were patients with ascending thoracic aortic aneurysms who were scheduled for elective surgery, transplant donors with normal aortas, and patients with acute aortic dissection. A subset of patients with ascending thoracic aortic aneurysms also had corresponding histological data. A separate subset of the ascending thoracic aortic aneurysm group underwent pre-operative MRI, and healthy volunteers without aortic disease underwent the identical MRI protocol as imaging controls. Written consent was obtained from all participants at the time of pre-operative clinic visits for elective aneurysm patients, post-operatively or through the substitute decision-maker in a deferred consent process for emergent dissection patients, or by research consent through Trillium Gift of Life for organ donors. A flow chart of encapsulating patients participating in the different phases of the study is shown in **Figure S1**.

### Tissue collection and preparation

Aortic tissue was collected from the operating room (or the multi-organ transplant coordinator in the case of tissue donors) and immediately placed on ice. Tissues were stored in Ringer’s lactate solution on ice during transportation (10-minute indoor walk from operating room to the laboratory) and kept at 4°C until testing. Mechanical testing was performed within 24 hours of resection as previously described^10^.

Ascending aortic tissue samples were collected as complete rings (**Figure 1a**) with the distal outer curvature marked with a surgical clip by the surgeon in the operating room. In the case of acute aortic dissections, the resected aortic tissue was also oriented by a clip with case-by-case explanation to the research team by the surgical team. The undissected portion, wherever that occurred for the aortic specimen, was used for biomechanical testing. Previous work by our group and others found that while some regional variability exists within a single ascending aorta, it is much less than variability between aortas, and that the biomechanics in one region of the aorta strongly predicted the biomechanics on the other side^42,43^.

**Fig. 1.**
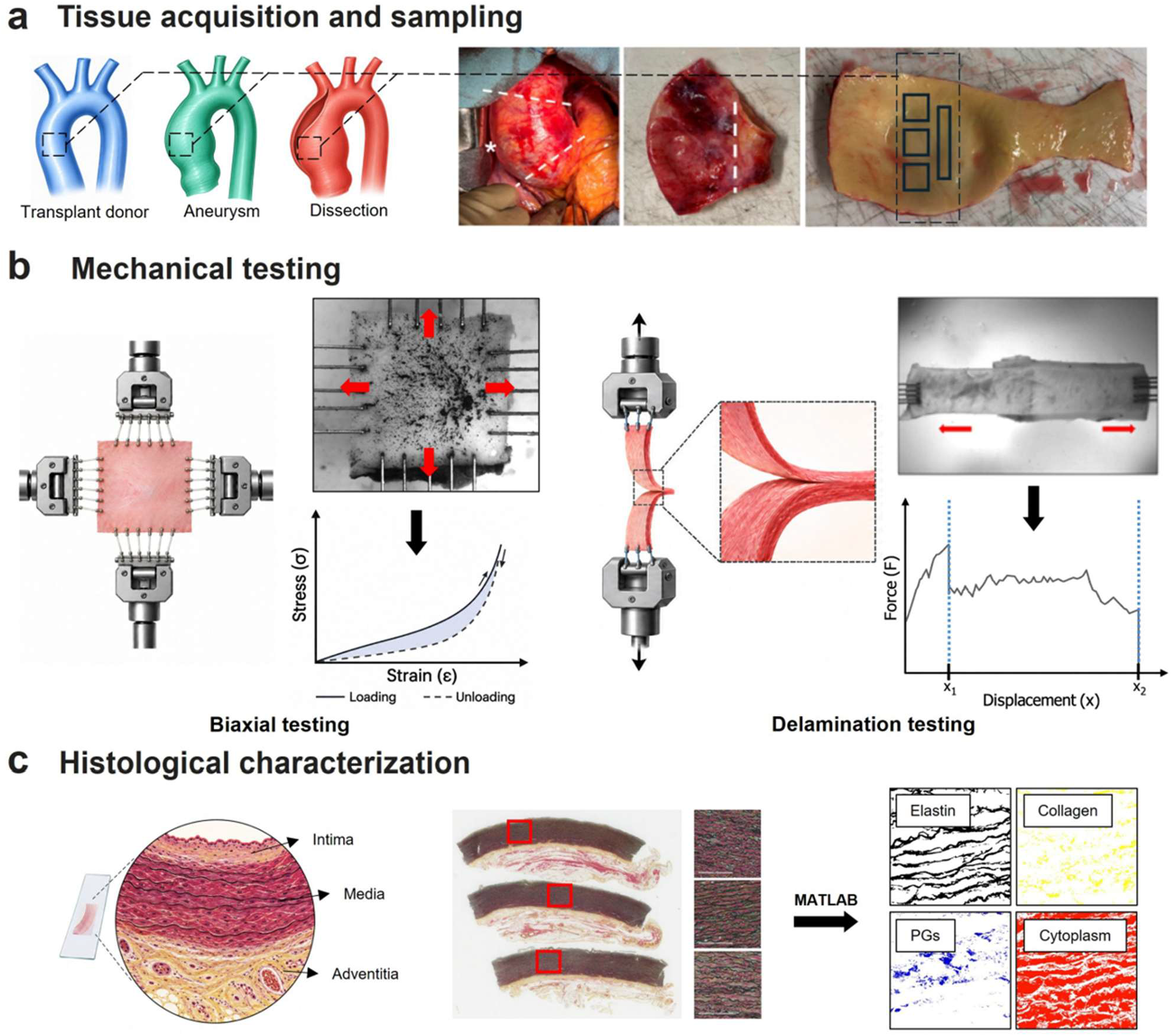
Tissue acquisition, mechanical testing and histological workflow. **a,** Ascending aortic tissue was collected as complete rings from the operating room, with regions marked for biaxial tensile testing and delamination (peel) testing. **b,** Schematic of biaxial tensile testing (left) and delamination testing (right), showing representative stress-strain and force-displacement curves, respectively, used to derive the low-strain modulus (LSM) and energy loss (EL) (biaxial test) and delamination strength (S_d_) (delamination test). **c,** Histological characterization workflow: Movat’s pentachrome-stained sections were used to quantify area fractions of elastin, collagen, proteoglycans (PGs) and smooth muscle cells (SMCs) by colorimetric segmentation.

From the outer curvature (OC), up to three 14 mm x 14 mm squares were sectioned for biaxial tensile testing. Additionally, ascending aortic tissue samples collected during surgery were cut into 6 mm x 30 mm strips from the OC adjacent to the samples taken for biaxial testing (**Figure 1b**).

### Biaxial tensile testing

The low strain modulus (LSM), defined as secant modulus from the origin to first deviation from linearity, and energy loss (EL) of the tissue specimens were measured through biaxial tensile testing using a BioTester (CellScale) as before^16,35,42^. First, tissue thickness was captured using a high magnification zoom lens (Navitar) and measured with ImageJ for subsequent stress calculations. Samples were then submerged in 37°C Ringer’s lactate solution and subject to 10 preconditioning cycles and 3 test cycles of 25% equibiaxial strain. Stress-strain curves generated from each of the three test cycles were then analyzed using an in-house MATLAB script to quantify the LSM and EL in the circumferential and longitudinal directions. Results from each test cycle were then averaged prior to use in subsequent analysis.

### Peel testing (delamination strength)

During aortic dissection, the aortic media splits along its length. The ease with which this failure mechanism may occur is represented by delamination strength (S_d_), measured as the average force required to peel the layers of the aortic wall normalized to the width of the delaminated sample. Tissues at higher risk of aortic dissection would exhibit lower delamination strengths^16^. The 6 mm x 30 mm strips obtained from the OC adjacent to the biaxial testing specimen were prepared by introducing a small incision into the medial layer using a scalpel. The tissue was peeled manually from proximal to distal end to leave 14 mm of intact tissue for testing. The strip was subsequently submerged in 37 °C Ringer’s lactate solution and subject to uniaxial peel testing using the BioTester^16,35,42,44^. Average force was calculated following initial peak (x1 in **Figure 1b**) and prior to complete failure (x2 in **Figure 1b**) to capture steady-state dissection propagation.

### Histology Analysis

Following biomechanical testing, the 14 × 14 mm outer-curvature square samples were fixed in 10% formalin and paraffin-embedded. Samples were cut circumferentially before edge-embedding to allow multiple regions of the tissue to be represented on each slide. Sections (4 μm) were stained with Movat’s pentachrome and digitally scanned at up to 20× magnification. Three 20× images of the aortic media were acquired at random locations from each slide. The area fractions of elastin, collagen, proteoglycans and cytoplasm were quantified by colorimetric analysis using previously published custom MATLAB scripts^45^, and the three measurements were averaged to obtain one value for each constituent per specimen. Histological analysis was limited to outer-curvature tissue and was not available for all specimens with complete mechanical testing.

### Pre-operative Magnetic Resonance Imaging (MRI)

Patients were imaged on a Siemens Magnetom Biograph mMR (3T) scanner using a dedicated 32-channel receiver coil. Standard 2D phase contrast (PC) images were acquired with breath holding and retrospective electrocardiographic (ECG)-gating at multiple defined locations, acquired perpendicular to the flow of blood at each level, sinotubular junction (STJ), mid-ascending aorta, and distal descending aorta at the level of the diaphragm (typical scan parameters 75-100 phases, 1.5 x 1.5 mm pixel spacing, 6 mm slice thickness, velocity encoding (VENC) of 150 cm/s). Balanced cine steady-state free precession (SSFP) imaging was obtained in sagittal oblique candy cane orientation for centerline length measurements.

### Derivation of in vivo biomechanical parameters

We investigated MRI derived markers for aortic stiffness including aortic strain, distensibility, compliance, and aortic stiffness index (ASI) as defined previously^19^. Aortic strain was defined as the change in cross-sectional area of the aorta divided by the minimal cross-sectional area. The related parameters of distensibility, compliance, and ASI were then derived by using maximal aortic strain and including the effect of pulsatile hemodynamic loading on the vessel wall as represented by the pulse pressure (difference between systolic and diastolic blood pressure)^46^. To measure the change in cross sectional area across the cardiac cycle, the aortic contours were segmented at mid-ascending aorta on 2D-PC MRI using CVI42 software, version 5.12.7 (Circle Cardiovascular Imaging, Canada). Each phase was reviewed by an operator and manual correction performed when the automatically detected boundaries deviated from the vessel wall. Standard systolic and diastolic brachial pressures by sphygmomanometer were obtained immediately prior to start of the MRI.

An alternative approach to estimate arterial stiffness independently of strain involves the measurement of pulse-wave velocity (PWV). PWV measures the speed of fluid wave propagation through a vessel and is related to the vessel stiffness by the Moens-Korteweg equation^47^. We measured aPWV using 2D-PC MRI acquired at the level of the STJ and the distal descending thoracic aorta (DDTA) at the aortic hiatus. The 2D-PC MRI contained encoded in-plane blood flow velocity that was used to a reconstruct blood flow over time curve at both STJ and DDTA levels using CVI42 software, version 5.12.7 (Circle Cardiovascular Imaging, Calgary, Canada). We then defined the zero of these flow curves as the arrival time of the fluid pulse wave and the lag between arrival times at STJ and DDTA as the transit time (TT) of the fluid pulse wave between both sampling sites. We measured the distance between both sampling sites as the centreline distance between STJ and DDTA on the three-dimensional (3D) reconstruction of the thoracic aorta. We then defined aPWV using Equation 1, where Δ*L* was the 3D centreline distance between sampling sites and TT was the transit time.

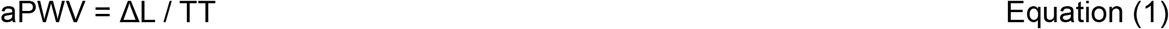

### Machine learning model for risk group classification based on ex vivo aortic biomechanics

The ground truth of how aortic biomechanics reflects risk of aortic dissection was defined by considering the ex vivo biomechanical properties of patients with aortic dissection and the normal aortas of transplant donors without aortic disease. To assess how the ex vivo properties jointly discriminate dissected from normal aortas, seven classifiers were evaluated including L2-logistic regression, elastic-net logistic regression, shrinkage linear discriminant analysis (shrinkage LDA), partial least square with logistic regression (PLS-LR), random forest, Gaussian process with a radial basis function kernel, and gradient boosted trees (XGBoost). Five ex vivo predictors were used including S_d_, longitudinal and circumferential EL, and longitudinal and circumferential LSM. To prevent information leakage, predictors were z-scored within each training fold using only that fold’s mean and standard deviation, applied as the first step of a pipeline fitted inside the cross-validation loop. Models were fitted by nested repeated stratified cross-validation with shuffling; the outer loop (five folds, five repeats, giving 455 out-of-fold predictions per model) provided out-of-fold predicted probabilities, while the inner loop (five folds, five repeats) selected the best hyperparameters by maximizing area under the receiver operating characteristic curve (AUC). Shrinkage LDA and the Gaussian process classifier required no inner search, since their regularization was calculated from the training fold directly by the Ledoit-Wolf shrinkage estimate and marginal likelihood respectively. Class weights were balanced wherever applicable. The pooled out-of-fold predictions were used to estimate discrimination performance, and the 95% confidence interval for the AUC was estimated using a 2,000-sample nonparametric bootstrap, with resampling performed at the patient level to preserve all repeated cross-validation predictions within each patient. Pairwise DeLong comparisons between classifiers used one repeat-averaged out-of-fold probability per patient (n = 91), rather than the pooled 455 predictions, to avoid treating correlated repeated cross-validation scores as independent observations. Discrimination was evaluated against a permutation-based null distribution. Class labels were permuted 1,000 times, and the full nested cross-validation procedure was refitted for each permutation using three outer repeats for computational tractability. The permutation *P* value was calculated using add-one smoothing, yielding a minimum attainable *P* value of 1/1,001. No post hoc recalibration was applied. Calibration of the raw predicted probabilities was assessed using the Brier score and the expected and maximum calibration errors across five equal-width probability bins, with confidence intervals obtained from the same patient-level bootstrap. From the out-of-fold predicted probabilities, two decision thresholds were defined to form a three-zone (“rule-out / intermediate / rule-in”) policy: (i) a low “rule-out” threshold set at the 5th percentile of the out-of-fold predicted probabilities among dissection patients, giving 95% sensitivity, and (ii) a high “rule-in” threshold set at the 95th percentile of the out-of-fold predicted probabilities among donor patients, giving 95% specificity. Because both thresholds were selected from the out-of-fold predictions, their operating characteristics were not considered independent estimates. Threshold selection was therefore repeated within cross-validation, with thresholds derived exclusively from each training fold’s out-of-fold predictions and then applied only to held-out patients who had no influence on their selection. After freezing the classifier, aneurysm cases were projected onto the risk classifier to visualize their position and classifier-estimated probability relative to the normal and dissection reference classes.

### Aortic Biomechanical vulnerability index (AortoBVI)

We refer to the continuous probability output of the classifier as the aortic biomechanical vulnerability index (AortoBVI). Values near 0 indicate a biomechanical profile similar to normal donor tissue, whereas values near 1 indicate a profile similar to tissue from patients with aortic dissection. The classifier was trained once using the donor and dissection specimens and was then fixed. AortoBVI values for elective aneurysm cases were therefore obtained by projecting them onto this fixed reference axis rather than by refitting the model. The aneurysm cases were subsequently positioned along this axis to show their relative similarity to the donor and dissection reference groups.

### Estimation of AortoBVI from MRI

Each aneurysm patient’s AortoBVI was obtained from the tissue-derived risk classifier. The regression target was then the logit-transformed AortoBVI, 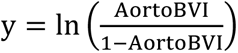, which maps AortoBVI’s bounded probability scale onto an unbounded continuous scale suitable for linear regression while preserving signed distance from the decision boundary. This target was modeled using ordinary least squares regression. To limit over-parameterization in this small cohort, the multivariable model family was prespecified to include age, aortic stiffness index (ASI), and pulse-wave velocity (aPWV). The strain-based family of in vivo biomechanical parameters (i.e., strain, distensibility, compliance, and ASI) are highly inter-related and derived from the same set of constituent variables^19^. Therefore, only ASI was selected as the representative pressure–deformation stiffness metric based on superior performance when comparing single-variable models. aPWV was included as a complementary velocity-based measure of aortic stiffening. Of candidate clinical parameters to include, our previous studies^16,48^ along with others^49–51^ have consistently reported age to be most predictive of ex vivo aortic biomechanics. Alongside the multivariable model and its single-predictor counterparts, maximum aortic diameter was also evaluated as a standalone benchmark, since it remains the primary parameter governing current clinical guidelines for risk stratification and intervention planning.

Candidate functional forms were intentionally restricted to linear terms to preserve interpretability and numerical stability at n=38. Because all models were fixed and prespecified in advance, no selection among equations was performed. Coefficient of determination (R^2^), root-mean-square error (RMSE), and mean absolute error (MAE) are reported both in the leave-one-out cross-validated (LOOCV) form and in-sample.

Each candidate equation generated a predicted AortoBVI for every aneurysm patient, representing the patient’s estimated location on the tissue-derived risk classifier. These predicted locations were then compared directly, on a patient-by-patient basis, against the corresponding tissue-derived AortoBVI from the frozen classifier. Uncertainty was characterized in terms of both precision and accuracy. Precision was quantified using a 95% prediction interval (PI) for each patient, incorporating residual variance and coefficient uncertainty. Prediction intervals were used rather than confidence intervals because the clinically relevant quantity is uncertainty in an individual patient prediction rather than in the mean response for patients with similar measurements. Because prediction intervals are model-specific, their widths were compared directly between models. Accuracy was assessed separately by determining whether each model’s prediction fell within a common tolerance of the measured value. A shared tolerance was required because evaluating each model against its own prediction interval would favour less precise models with wider intervals. The tolerance was therefore fixed at the smallest cross-validated RMSE of all models since fixing it at the larger value would have let both models clear a lenient band and obscured any real difference between them. Model performance was assessed by examining how well these predicted locations captured the true patient locations, including with respect to the predefined rule-out, intermediate zone, and rule-in boundaries.

### Statistical analysis

All statistical tests were two-sided, and the patient was the unit of analysis throughout. Repeated biaxial test cycles, adjacent outer-curvature tissue regions and the three histological fields acquired per slide were averaged to obtain a single patient-level value before statistical analysis. Continuous variables are reported as mean ± s.d., except aortic diameter, which is reported as median [interquartile range], and categorical variables as n/N (%) or n (%) as appropriate. Normality was not assumed, and non-parametric tests were used for group comparisons because several ex vivo mechanical and MRI-derived variables showed non-Gaussian distributions. Differences among transplant donor, aneurysm and dissection groups were assessed using Kruskal–Wallis tests. Maximum diameter was compared between aneurysm and dissection groups using a two-sided Mann–Whitney test. Omnibus *P* values were Holm-corrected across the five prespecified ex vivo mechanical comparisons, and pairwise group differences were assessed using Dunn’s post-hoc tests with Holm correction. Categorical variables were compared using the chi-square test or, when any expected cell count was below five, the Fisher–Freeman–Halton test with Monte Carlo estimation of the *P* value (20,000 replicates). For comparisons between aneurysm patients and healthy MRI volunteers, continuous variables were assessed using two-sided Mann–Whitney tests and categorical variables using two-sided Fisher’s exact tests. *P* values for the five MRI-derived variables were Holm-corrected across comparisons. Associations between the five MRI-derived indices and five ex vivo mechanical properties were first assessed using ordinary linear regression. Inference across this prespecified family of 25 MRI–mechanics associations used HC3 heteroskedasticity-robust standard errors, with *P* values adjusted across all 25 tests using the Benjamini–Hochberg procedure and reported as *q* values. Spearman rank correlations, with Benjamini–Hochberg correction across the same 25 pairs, were additionally calculated as a rank-based sensitivity analysis. We further modelled the relationship for some in vivo parameters (strain, distensibility, compliance, and ASI) using piecewise linear regression due to noted nonlinearity. The breakpoint was estimated as the value jointly maximizing the summed *R*² across all five ex vivo outcomes, with breakpoints constrained to values supported by at least 20% of patients on each side. This criterion was met on the raw scale for strain, distensibility and compliance. For ASI, because of its more heavily right-skewed distribution, adequate support on both sides was achieved only on the natural log scale. The resulting breakpoint was therefore back-transformed to raw units and modelled as a linear breakpoint on the raw scale, consistent with the other three parameters. The breakpoint was additionally tested with Davies test in Python using the ‘piece-wise regression’ toolbox^52^. For estimation of tissue-derived AortoBVI from MRI, predictive performance was evaluated using leave-one-out cross-validation (LOOCV) and summarized by cross-validated *R*^2^, root-mean-square error (RMSE) and mean absolute error (MAE), together with Pearson r and Lin’s concordance correlation coefficient. Apparent in-sample *R*^2^ values were reported separately for comparison. Histological constituents were compared across the three AortoBVI zones using Kruskal–Wallis tests with Dunn’s post-hoc tests, and correlated with continuous tissue-derived logit AortoBVI using Spearman rank correlation, with Benjamini–Hochberg correction applied across the five constituents. Associations between each constituent and AortoBVI were further assessed using sequential linear regression models with HC3 heteroskedasticity-robust standard errors, adjusting stepwise for age, diameter, sex and bicuspid aortic valve status, with standardized coefficients reported and *P* values Benjamini–Hochberg-corrected across the five constituents within each adjustment level. A selection audit compared patients with and without available histology using standardized mean differences for tissue-derived AortoBVI and the five ex vivo mechanical parameters. Statistical significance was defined as two-sided *P* < 0.05 or multiplicity-adjusted P or q < 0.05, as applicable. Analyses were performed in Python 3.13 using NumPy 2.5, pandas 3.0, SciPy 1.18, scikit-learn 1.9, statsmodels 0.14.6, scikit-posthocs 0.14, xgboost 3.4 and piecewise-regression 1.5. Random seeds were fixed for all stochastic procedures. Statistical significance was defined as two-sided *P* < 0.05 or multiplicity-adjusted *P* or *q* < 0.05, as applicable.

## Results

### Ex vivo biomechanics differed across normal, aneurysm, and dissected aortas

For the ex vivo biomechanical analysis, 353 elective ascending thoracic aortic aneurysm (ATAA) patients, 45 transplant donors with normal aortas, and 46 patients with acute aortic dissection underwent biaxial tensile and delamination testing **(Figure 1 and Figure S1)**. Baseline characteristics are summarized in **Table 1**. In the ATAA group, the mean age was 61.3 ± 13.5 years, 252 patients (71.4%) were male, 143 patients (40.5%) had a bicuspid aortic valve, and the median maximum aortic diameter was 52.0 [48.0–55.0] mm. In the donor group, the mean age was 46.4 ± 13.8 years, and 28 patients (65.1%) were male; aortic diameter was not recorded because preoperative imaging was unavailable. In the dissection group, the mean age was 65.8 ± 14.1 years, 31 patients (67.4%) were male, and the median maximum aortic diameter was 48.5 [45.0–53.8] mm.

**Table 1.**
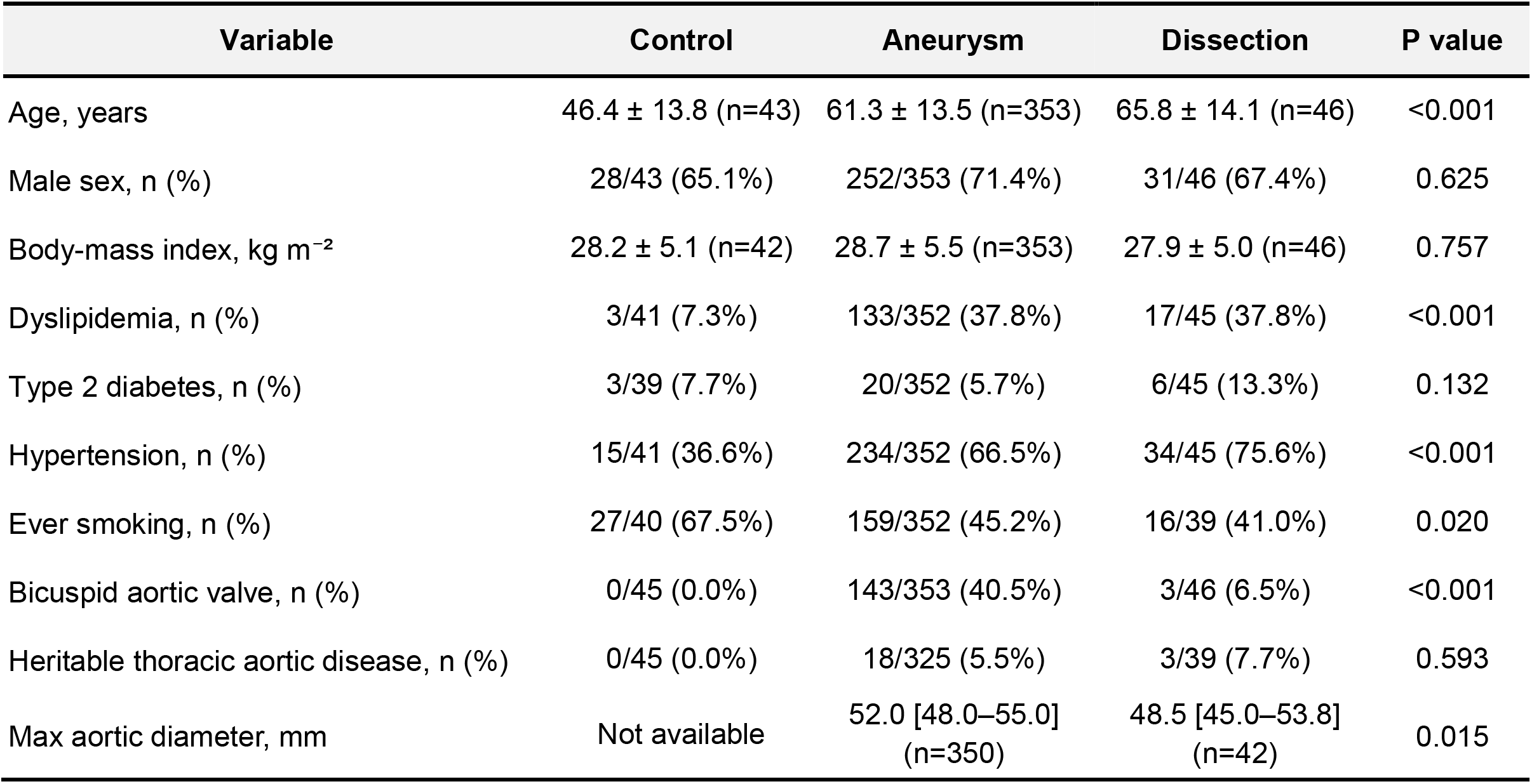
Clinical characteristics of the complete-mechanics tissue cohort. Demographic and clinical characteristics of transplant donors with normal aortas and patients with elective aneurysm or acute type A dissection with complete ex vivo mechanical testing. Continuous variables are reported as mean ± s.d., except maximum aortic diameter, which is reported as median [interquartile range]; categorical variables are reported as n (%). Group differences were assessed using the Kruskal–Wallis test for continuous variables and the chi-square or Fisher–Freeman–Halton test for categorical variables, as appropriate. Diameter is compared between aneurysm and dissection groups using the Mann–Whitney test.

| Variable | Control | Aneurysm | Dissection | P value |
|---|---|---|---|---|
| Age, years | 46.4 ± 13.8 (n=43) | 61.3 ± 13.5 (n=353) | 65.8 ± 14.1 (n=46) | <0.001 |
| Male sex, n (%) | 28/43 (65.1%) | 252/353 (71.4%) | 31/46 (67.4%) | 0.625 |
| Body-mass index, kg m<sup>−2</sup> | 28.2 ± 5.1 (n=42) | 28.7 ± 5.5 (n=353) | 27.9 ± 5.0 (n=46) | 0.757 |
| Dyslipidemia, n (%) | 3/41 (7.3%) | 133/352 (37.8%) | 17/45 (37.8%) | <0.001 |
| Type 2 diabetes, n (%) | 3/39 (7.7%) | 20/352 (5.7%) | 6/45 (13.3%) | 0.132 |
| Hypertension, n (%) | 15/41 (36.6%) | 234/352 (66.5%) | 34/45 (75.6%) | <0.001 |
| Ever smoking, n (%) | 27/40 (67.5%) | 159/352 (45.2%) | 16/39 (41.0%) | 0.020 |
| Bicuspid aortic valve, n (%) | 0/45 (0.0%) | 143/353 (40.5%) | 3/46 (6.5%) | <0.001 |
| Heritable thoracic aortic disease, n (%) | 0/45 (0.0%) | 18/325 (5.5%) | 3/39 (7.7%) | 0.593 |
| Max aortic diameter, mm | Not available | 52.0 [48.0–55.0] (n=350) | 48.5 [45.0–53.8] (n=42) | 0.015 |

All ex vivo biomechanical properties differed among transplant donors, ATAA, and dissection groups after Holm correction across the five Kruskal-Wallis tests (all H(2) ≥ 12.41, adjusted *P* ≤ 0.002; **Figure 2**). Circumferential EL (EL_circ_) was significantly higher in aneurysm and dissection patients than in donors (**Figure 2a**). In contrast, circumferential LSM (LSM_circ_) was lower in dissection patients than in donors (*P* = 0.011, **Figure 2b**) and aneurysm patients (*P* < 0.001, **Figure 2b**). Longitudinal EL (EL_long_) showed a similar pattern to EL_circ_, being higher in both aneurysm and dissection patients than in donors (P < 0.001; **Figure 2c**). Longitudinal LSM (LSM_long_) was lower in dissection than in aneurysm patients (P = 0.001; **Figure 2d**), but did not differ significantly between dissection patients and donors (P = 0.095). Delamination strength (S_d_) was lower in aneurysm and dissection patients than in donors (both *P* < 0.001, **Figure 2e**) and in dissection than aneurysm patients (*P* = 0.013, **Figure 2e**).

**Fig. 2.**
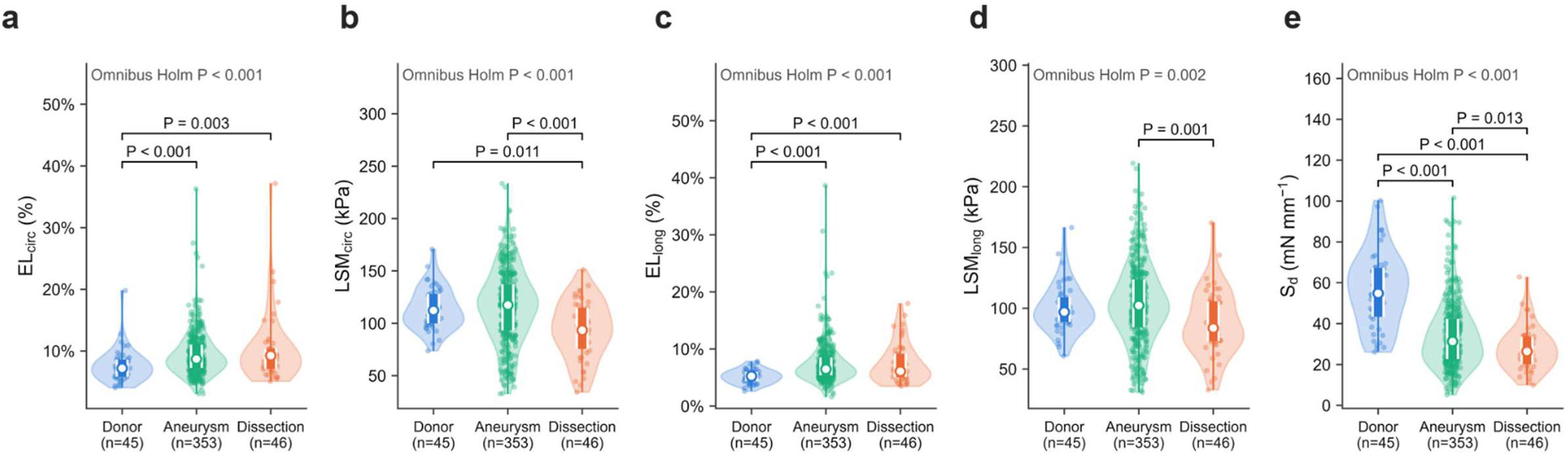
Mechanical properties differ across tissue groups. Violin plots with overlaid box plots and individual observations showing circumferential energy loss (**a**), circumferential low-strain modulus (**b**), longitudinal energy loss (**c**), longitudinal low-strain modulus (**d**), and delamination strength (**e**) in transplant donor with normal aorta (n = 45), aneurysm (n = 353) and dissection (n = 46) tissues. Omnibus P values were calculated using Kruskal–Wallis tests and Holm-corrected across all prespecified ex vivo mechanical comparisons. Pairwise *P* values were calculated using Dunn’s post-hoc tests with Holm correction. White circles indicate medians and boxes indicate interquartile ranges.

### A classifier separated normal from dissected aortas

We trained seven supervised binary classifiers to separate donor (N = 45) from dissected aortas (N = 46) using five ex vivo biomechanical parameters. Across all classifiers, discrimination was comparable (out-of-fold AUCs = 0.874-0.915; **Figure 3a**). Bootstrap confidence intervals showed substantial overlap across models (**Figure 3b**), with no significant pairwise differences after multiplicity correction of the DeLong tests (minimum adjusted *q* = 0.175). The full seven-model comparison is provided in **Tables S1a and S1b**.

**Fig. 3.**
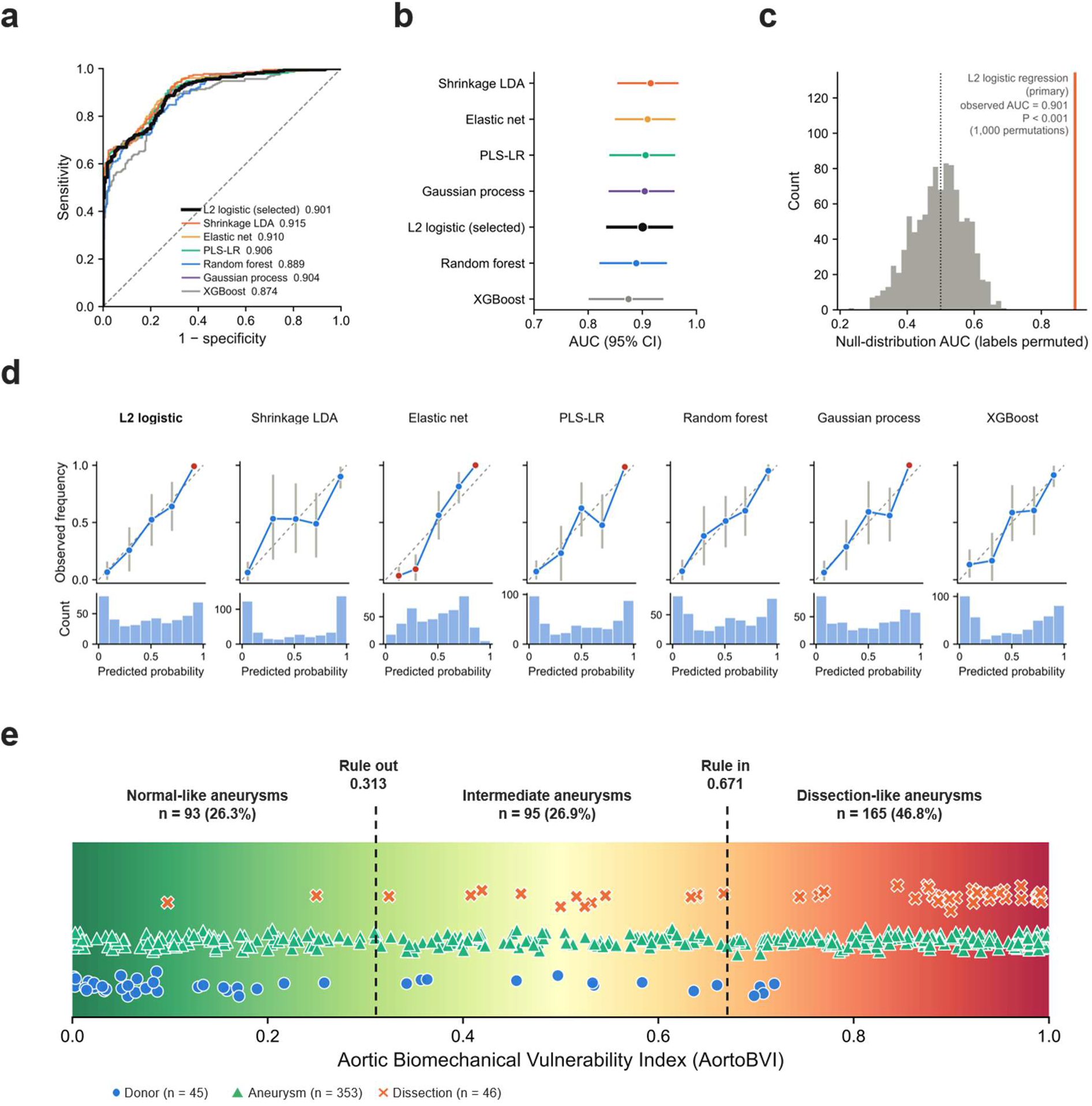
A frozen tissue classifier defines a continuous biomechanical phenotype. **a**, Receiver operating characteristic (ROC) curves for seven candidate classifiers trained to distinguish donor (n = 45) from dissection (n = 46) tissue using five ex vivo mechanical predictors; the primary L2-regularized logistic classifier is highlighted. **b,** Comparison of out-of-fold AUCs across the seven candidate classifiers; points indicate AUC estimates and horizontal bars indicate 95% confidence intervals. **c,** Null distribution of AUCs obtained from 1,000 label permutations for the primary L2-regularized logistic classifier, with the observed out-of-fold AUC indicated by the vertical orange line; the dotted line indicates the mean null AUC (*P* < 0.001). **d,** Calibration of predicted probabilities for each candidate classifier. Upper panels show observed event frequency versus predicted probability across probability bins, with the dashed diagonal indicating perfect calibration; lower panels show the corresponding distributions of predicted probabilities. E, Aortic Biomechanical Vulnerability Index (AortoBVI) values for 353 elective aneurysm patients, stratified into normal-like, intermediate and dissection-like zones using rule-out (AortoBVI = 0.313) and rule-in (AortoBVI = 0.671) thresholds derived from out-of-fold classifier predictions. Points within each group are vertically jittered for visual separation; y position has no quantitative meaning.

Among classifiers without statistically significant pairwise differences, we selected L2-regularized logistic classifier as the primary model for its clinical familiarity and closed-form deployment equation. This classifier distinguished donors from dissected aortas with strong discriminative performance (nested cross-validation out-of-fold AUC = 0.901, 95% bootstrap CI 0.836–0.954). Permutation testing with 1,000 label permutations confirmed that this discrimination exceeded that expected by chance (*P* < 0.001; mean null AUC = 0.495; **Figure 3c**). The model also showed good calibration, with a Brier score of 0.129 and an expected calibration error of 0.042 (**Figure 3d and Tables S1a**).

### Aortic biomechanical vulnerability index (AortoBVI) stratified patients into clinically meaningful categories

The fitted L2 classifier’s continuous probability output was defined as the Aortic Biomechanical Vulnerability Index (AortoBVI) and calculated for all 353 elective aneurysms without model refitting. To translate AortoBVI into clinically interpretable risk categories, we defined two clinically motivated thresholds forming a three-zone risk classifier: a rule-out threshold (threshold_low_ = 0.313) corresponding to 95% sensitivity and a negative predictive value of 0.909, and a rule-in threshold (threshold_high_ = 0.671) corresponding to 95% specificity, yielding a positive predictive value of 0.912. Between these thresholds, cases were classified as an “intermediate zone” phenotype. Aneurysm patients spanned across these decision boundaries, with 26.3% (N = 93) falling below the rule-out threshold (normal-like biomechanical phenotype), 46.8% (N = 165) exceeding the rule-in threshold (dissection-like phenotype), and 26.9% (N = 95) lying within the intermediate risk zone **(Figure 3e).** A parallel sensitivity/specificity analysis and threshold re-selection altered classification proportions for three alternative boundary definitions (**Figure S2)**.

### AortoBVI correlated with extracellular matrix content

As an independent biological validation of the biomechanics-derived phenotypes, we compared histology across the three AortoBVI zones in 184 aneurysm patients with available histology (**Figure 1c**). The histology subset was representative of the full cohort with respect to AortoBVI (standardized mean difference = 0.011; **Figure S3a**). Elastin area fraction differed across zones (Kruskal–Wallis *P* < 0.001, **Figure 4a**), with lower elastin in dissection-like than normal-like and intermediate patients (both *P* < 0.001). Collagen area fraction also differed across zones (Kruskal–Wallis *P* = 0.007, **Figure 4b**), with dissection-like patients showing higher collagen content than normal-like patients (*P* = 0.006). As a continuous measure, tissue-derived AortoBVI correlated inversely with elastin (Spearman ρ = -0.458, q < 0.001) and positively with collagen (ρ = 0.288, q < 0.001) (**Figure S3b,c**). The elastin association persisted after sequential adjustment for age, diameter, sex and bicuspid aortic valve status (fully adjusted standardized β = −0.303; q = 0.013), whereas the collagen association did not survive after adjustment for age and was not significant in the fully adjusted model (q = 0.179; **Figure S3d**).

**Fig. 4.**
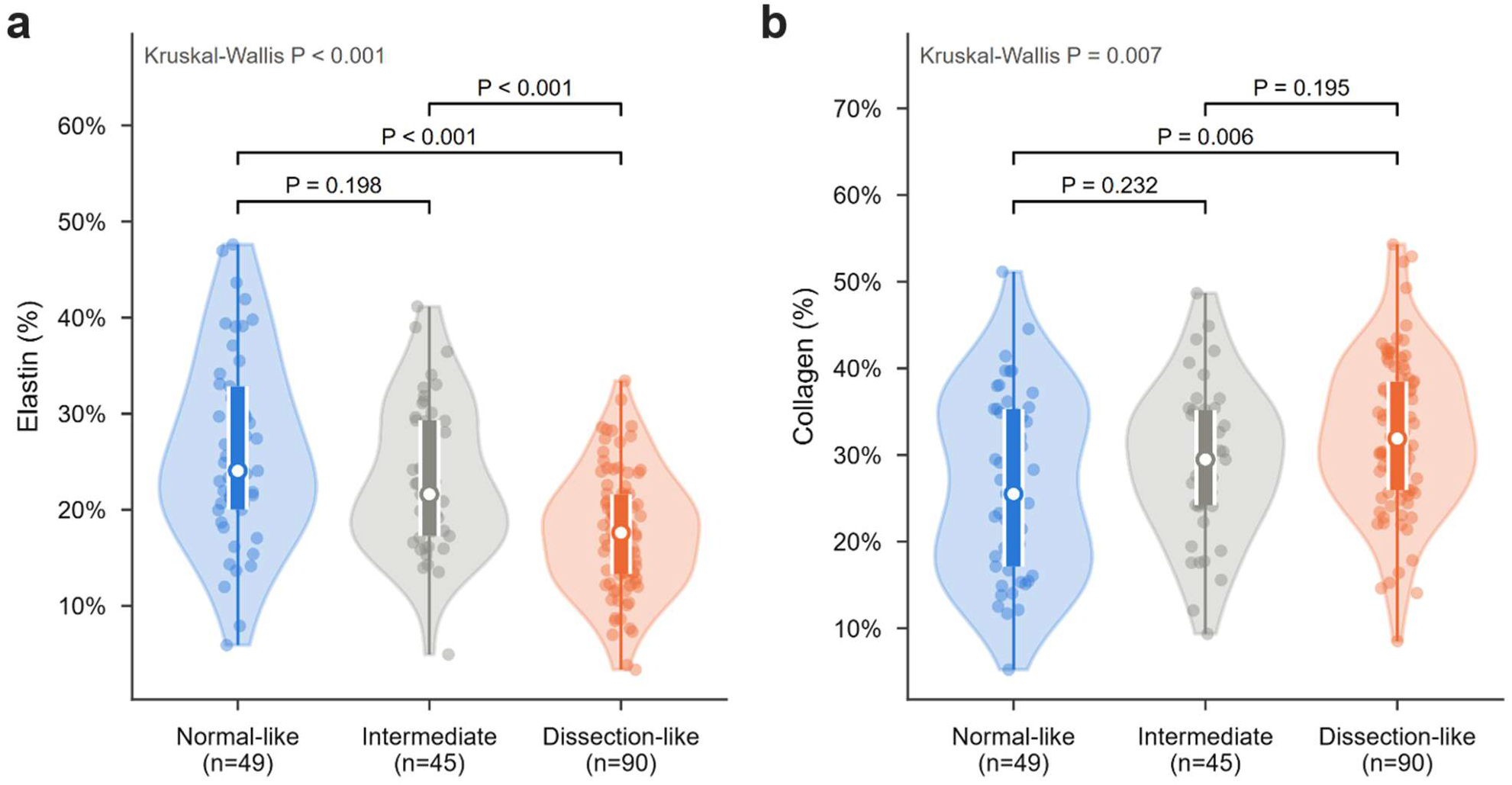
Histological components ofthe aortic biomechanical phenotypes. Violin plots showing elastin (**a**) and collagen (**b**) area fractions across AortoBVI zones (normal-like, n = 49; intermediate, n = 45; dissection-like, n = 90). Omnibus *P* values were calculated using Kruskal–Wallis tests, and pairwise *P* values using Dunn’s post-hoc tests with Holm correction.

### MRI-derived parameters did not differ between ATAA patients and healthy controls

For AortoBVI to be useful for clinical risk assessment, it must be predictable non-invasively using, e.g, medical imaging from which in vivo biomechanical parameters can be estimated. Among ATAA patients whose aortas underwent mechanical testing, 44 had preoperative MRI with at least one MRI-derived biomechanical parameter (strain, distensibility, compliance, ASI, or aPWV) available. Of these, 38 had complete MRI data and corresponding tissue measurements and were included in the paired in vivo–ex vivo analyses. An additional 8 healthy volunteers also underwent the same MRI protocol **(Figure S4)**. Baseline characteristics of the MRI-available cohorts are summarized in **Table 2**.

**Table 2.** Clinical characteristics of the MRI-available cohort. Demographic and clinical characteristics of healthy MRI volunteers and patients with aneurysm who underwent preoperative MRI. Continuous variables are reported as mean ± s.d., except maximum aortic diameter, which is reported as median [interquartile range]; categorical variables are reported as n (%). Group differences were assessed using two-sided Mann–Whitney tests for continuous variables and two-sided Fisher’s exact tests for categorical variables.

| **Variable** | **Healthy (N=8)** | **Aneurysm (N=44)** | **P value** |
|---|---|---|---|
| Age, years | 52.1 ± 16.1 (n=8) | 59.8 ± 12.9 (n=44) | 0.125 |
| Body-mass index, kg m<sup>−2</sup> | 24.5 ± 3.3 (n=6) | 28.7 ± 6.0 (n=44) | 0.018 |
| Male sex, n/N (%) | 3/8 (37.5%) | 36/44 (81.8%) | 0.017 |
| Dyslipidemia, n/N (%) | 0/7 (0.0%) | 14/44 (31.8%) | 0.169 |
| Type 2 diabetes, n/N (%) | 0/7 (0.0%) | 0/44 (0.0%) | 1.0 |
| Hypertension, n/N (%) | 2/7 (28.6%) | 28/44 (63.6%) | 0.109 |
| Ever smoking, n/N (%) | 0/7 (0.0%) | 15/44 (34.1%) | 0.090 |
| Max aortic diameter, mm | 30.5 [28.8–32.5] (n=8) | 52.0 [50.0–54.2] (n=44) | <0.001 |

We first tested if individual MRI-derived biomechanical parameters differed between normal and diseased states, but none of the five measured parameters differed significantly between ATAA patients and healthy imaging controls after Holm correction (Holm *P* > 0.125; **Figure S4**).

### ASI associated most consistently with ex vivo biomechanics

We next considered relationships between in vivo MRI-derived vs. ex vivo biomechanical parameters (**Figure 5**). Linear regression yielded some significant associations (*q* < 0.05), most notably ASI with all ex vivo parameters, but with weak to moderate R² of 0.20 to 0.31 (**Figure S5).** Because several in vivo–ex vivo relationships exhibited inflection points not captured by a linear model, we used piecewise linear regression to assess whether model fit improved (**Figure 5 and Figure S6).** Piecewise regression comparing aortic strain with the circumferential metrics yielded R² = 0.29 for EL_circ_, R² = 0.076 for LSM_circ_, and R² = 0.23 for S_d_, all improved over the corresponding linear models (**Figure 5a–c and Figure S5**). The piecewise model identified a breakpoint at 7.20% strain. Overall, higher strain was associated with lower EL_circ_ and higher S_d_, while the relationship with LSM_circ_ was not significant. The corresponding piecewise relationships for the longitudinal metrics also follow similar patterns with R² = 0.21 for EL_long_ and R² = 0.14 for LSM_long_ **(Figure S6a–b)**. Similarly, piecewise regression for distensibility improved over the linear models with R^2^ = 0.30 for EL_circ_, R^2^ = 0.22 for LSM_circ_, and R^2^ = 0.24 for S_d_, with a breakpoint at 1.19 × 10^−3^ mmHg^-1^ (**Figure 5d–f**). The corresponding longitudinal relationships yielded R² = 0.34 for EL_long_ and R² = 0.28 for LSM_long_ (**Figure S6c–d**). For compliance, the piecewise models yielded R^2^ = 0.25 for EL_circ_, R^2^ = 0.17 for LSM_circ_, and R^2^= 0.087 for S_d_ with the breakpoint at 1.77 mm² mmHg^−1^ (**Figure 5g–i**). Longitudinal fits yielded R^2^ = 0.23 for EL_long_ and R^2^ = 0.24 LSM_long_ (**Figure S6e–f**). For ASI, the piecewise models improved over the linear models and were significantly associated with all five ex vivo parameters, with R^2^ = 0.23 for EL_circ_, R^2^ = 0.36 for LSM_circ_, and R^2^ = 0.20 for S_d_, with a breakpoint at 9.27 (**Figure 5j–l**). Longitudinal relationships showed R^2^ = 0.31 for EL_long_ and R^2^ = 0.34 for LSM_long_ (**Figure S6g–h**). In contrast to the strain-derived parameters, aPWV showed no significant association with ex vivo biomechanics in either direction (R² = 0.005–0.060, all *P* ≥ 0.139), and no breakpoint was detected (**Figure 5m–o and Figure S6i–j**).

**Fig. 5.**
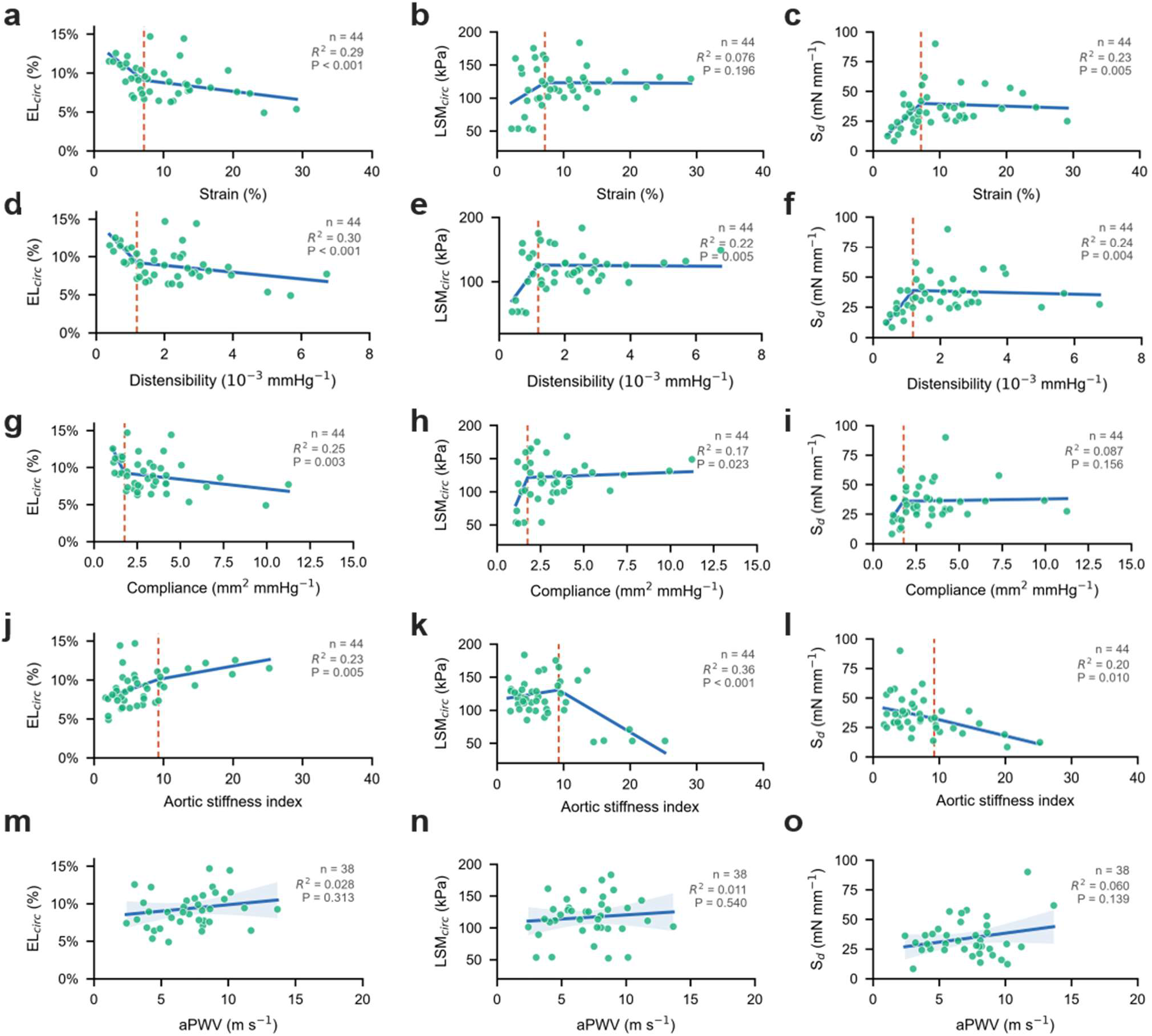
MRI-derived indices are associated with ex vivo tissue mechanics. Piecewise linear regressions of circumferential energy loss (left column), circumferential low-strain modulus (middle column) and delamination strength (right column) against maximal aortic strain **(a–c)**, distensibility **(d–f)**, compliance **(g–i)** and aortic stiffness index **(j–l)**, and simple linear regressions against aortic pulse-wave velocity **(m–o)**. Dashed vertical lines indicate estimated breakpoints for strain, distensibility, compliance and aortic stiffness index. Breakpoint significance was assessed using the Davies test. *R²* and *P* values from the overall regression model are shown in each panel (n = 44 for strain, distensibility, compliance and aortic stiffness index; n = 38 for aortic pulse-wave velocity).

### MRI-derived measures estimated AortoBVI more closely than diameter

Given the strong performance of AortoBVI in stratifying patients by biomechanical phenotypes, we used AortoBVI to evaluate candidate imaging models. For the 38 aneurysm patients with complete imaging and tissue measurements, AortoBVI classified 8 patients (21.1%) as normal-like, 15 patients (39.5%) as dissection-like, and 15 (39.5%) patients as intermediate.

We evaluated five prespecified imaging models against tissue-derived AortoBVI using leave-one-out cross-validation (**Figure 6a**): ASI alone, age alone, aPWV alone, maximum aortic diameter alone, and a multivariable model combining age, ASI and aPWV. Age was included for its established association with ex vivo biomechanics^16^, ASI for its strong and consistent associations with ex vivo biomechanics among strain-derived indices (**Figure 5)**, and aPWV as a complementary velocity-based measure of aortic stiffening. Diameter was deliberately excluded from the multivariable model and evaluated separately as the clinical benchmark, allowing direct comparison with current intervention criteria.

**Fig. 6.**
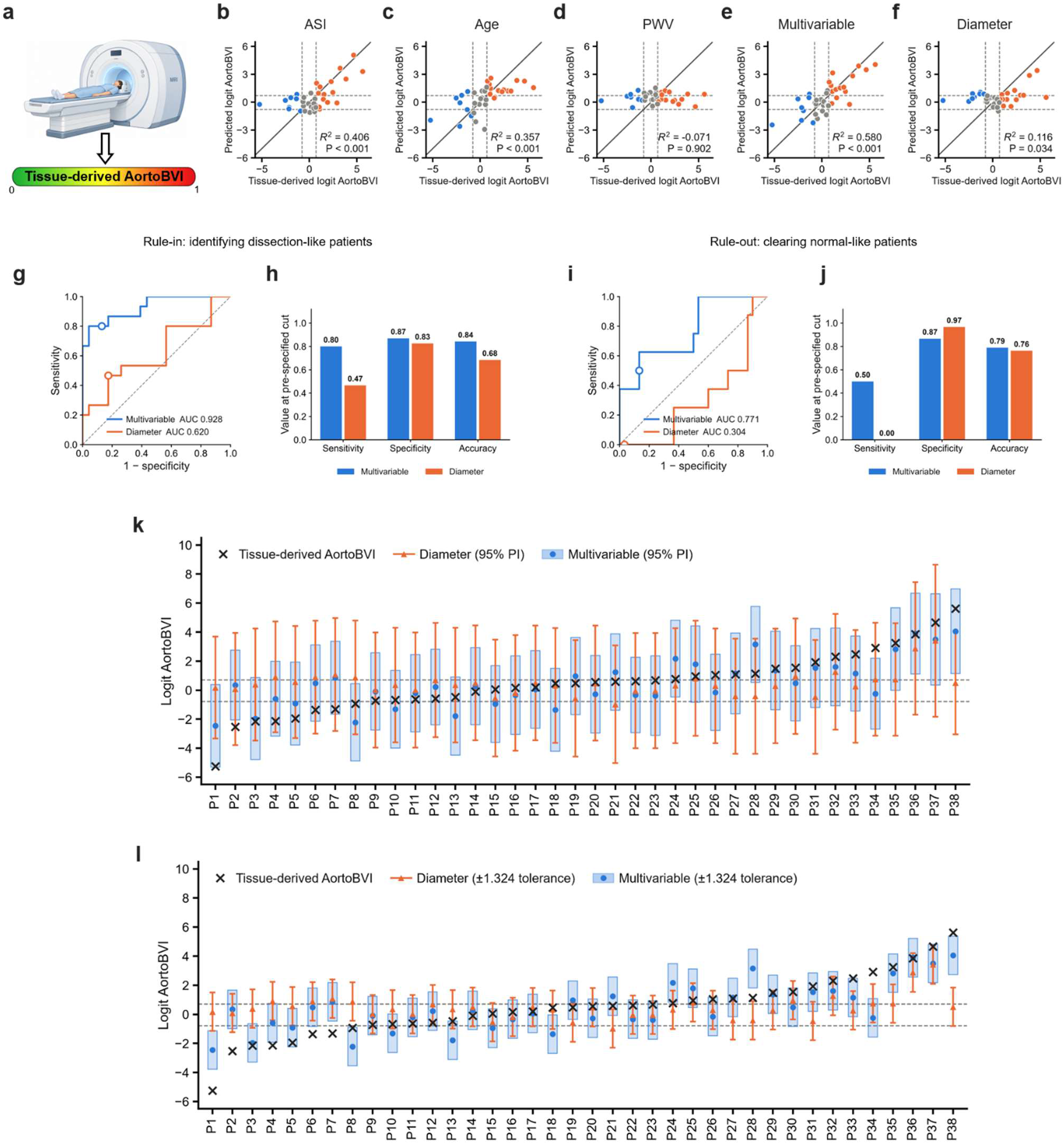
Five prespecified imaging models estimate tissue-derived AortoBVI. **a**, Schematic illustrating prediction of tissue-derived AortoBVI from non-invasive imaging and clinical measurements. **b–f,** Leave-one-out cross-validated predicted versus tissue-derived logit-transformed AortoBVI for models based on **b,** ASI alone, **c,** age alone, **d,** aPWV alone, **e,** a multivariable model combining age, ASI, and aPWV, **f**, and maximum aortic diameter alone. Points are coloured by BVI zone; leave-one-out cross-validated *R*² and *P* values are shown for each model (n = 38). **g, h,** Rule-in analysis distinguishing dissection-like cases (n = 15) from the remaining cohort (n = 23). Receiver operating characteristic curves **(g)** and operating characteristics at the selected threshold **(h)** comparing the multivariable model with diameter alone. **i, j,** Rule-out analysis distinguishing normal-like cases (n = 8) from the remaining cohort (n = 30). Receiver operating characteristic curves **(i)** and operating characteristics at the selected threshold **(j)** comparing the multivariable model with diameter alone. **k**, Patient-level predictions from the diameter-only and multivariable models, with model-specific 95% prediction intervals, ordered by tissue-derived AortoBVI. **l,** Patient-level prediction errors assessed against a common tolerance band defined by the smallest cross-validated RMSE across the five models.

The full comparison of all five models is shown in **Figure 6** and **Table S2**. ASI was a stronger single predictor of tissue-derived AortoBVI (*R*² = 0.406, *P* < 0.001; **Figure 6b**) than age (*R*² = 0.357, *P* < 0.001; **Figure 6c**) or aPWV (not significant; **Figure 6d)**, whereas the multivariable model achieved the best overall performance (*R*² = 0.580, *P* < 0.001; **Figure 6e**), exceeding diameter alone which was weakly (*R*² = 0.116, *P*=0.034) correlated with AortoBVI (**Figure 6f**). This represented nearly a 5-fold increase in explained variation (58.0% versus 11.6%). The multivariable model also had lower prediction error than diameter (RMSE, 1.324 versus 1.921; MAE, 1.075 versus 1.497) and stronger agreement with tissue-derived AortoBVI (Lin’s concordance correlation coefficient, 0.742 versus 0.236). Additionally, for distinguishing the dissection-like zone (n = 15) from the remainder of the cohort (normal-like and intermediate, n = 23) at the rule-in threshold (AortoBVI = 0.671), the multivariable model achieved an AUC of 0.928 (95% CI, 0.826–0.994; **Figure 6g**), with 80% sensitivity, 87% specificity and 84% accuracy (**Figure 6h**), whereas diameter alone showed limited discrimination (AUC = 0.620; Figure **6g**), with 47% sensitivity, 83% specificity and 68% accuracy (**Figure 6h**). For distinguishing the normal-like zone (n = 8) from the remainder of the cohort (intermediate and dissection-like, n = 30) at the rule-out threshold (AortoBVI = 0.313), the multivariable model again showed greater discrimination than diameter (AUC = 0.771 versus 0.304; **Figure 6i**), identifying 50% of normal-like patients with 87% specificity and 79% accuracy, compared with 0% sensitivity for diameter despite its higher specificity (97%) and similar overall accuracy (76%) (**Figure 6j**). The higher specificity of diameter in this analysis resulted from its failure to classify any patients as normal-like, rather than from accurate identification of the normal-like phenotype (0% sensitivity). At the individual-patient level, the 95% prediction intervals for the multivariable model were narrower in all patients than those of diameter, with a 32.1% mean reduction in width (**Figure 6k**). Using a common tolerance of ±1.324 logit-AortoBVI units, the lowest cross-validated RMSE across all models, 27 of 38 multivariable predictions fell within tolerance of the tissue-derived AortoBVI, compared with 22 of 38 diameter predictions (**Figure 6l**). Exact three-zone agreement was also better for the multivariable model (25 of 38 patients, 65.8%, **Figure S7a**) than for diameter alone (21 of 38, 55.3%, **Figure S7b**). Although neither model classified a dissection-like patient as normal-like, diameter classified 4 of 8 normal-like patients as dissection-like, compared with 1 of 8 for the multivariable model (**Figure S7a, b)**. Because ASI was the strongest MRI-derived predictor, we further examined whether a simple ASI threshold could enrich for the dissection-like phenotype. An ASI threshold >9.49 was exceeded exclusively by dissection-like patients, identifying 9 of 15 cases (60%) without any normal-like or intermediate cases exceeding this value (**Figure S7c**). This threshold was close to the breakpoint independently identified in the ex vivo piecewise analysis (ASI = 9.27; **Figure 5j–l**), despite being derived from a different cohort and a different criterion.

Additionally, as a face-validity check, the multivariable model classified eight independent healthy MRI volunteers as five normal-like (AortoBVI 0.189 ± 0.072) and three intermediate (AortoBVI 0.471 ± 0.076), with no dissection-like. The three intermediate volunteers were aged 45, 67 and 78 years (AortoBVI 0.384, 0.515 and 0.515) (**Figure S7d**).

## Discussion

Biomechanics of the ascending aorta are nonlinear and anisotropic, driven by the interplay among extracellular matrix components^18,29–31^. In the aneurysmal state, aortic stress-strain behaviour is influenced by the heterogeneous nature of medial degeneration. Elastin fragmentation and loss, lamellar unit collapse, accumulation of immature disorganized collagen and pooling of glycosaminoglycans vary across diseased aortas and jointly influence aortic mechanical behaviour at low and high stresses, the transitions between stress domains, and aortic viscoelastic behaviour^13,18,30^. These complexities have prevented any single mechanical metric from capturing aortic wall fragility, which clinically manifests as aortic dissection, rupture and death. Aortic diameter, the dominant clinical metric, is even further removed, acting as an indirect surrogate of wall stress and disease severity, and likewise fails to capture aortic wall fragility.

To address this need, we avoided reductionism and integrated multiple biomechanical parameters via a machine learning approach capable of handling correlated variables, reflecting the nonlinear, heterogeneous nature of aortic wall mechanical behavior. The key innovation was to connect aortic biomechanics to clinically relevant endpoints and then present a pathway to clinical translation via imaging. Previous attempts to evaluate determinants of aortic risk have been hampered by the rarity of aortic dissection^21,32,33^, limiting traditional epidemiological approaches such as prospective trials. Alternative unproven endpoints such as growth are sometimes used^21,32,33^, but these studies are muddied by surgical interventions curtailing natural history^34^. The advent of machine learning in medicine has allowed this problem to be reimagined. By uniquely curating dissected and normal aortas (transplant donors) over 10 years, we trained our model on patients with unambiguous risk categorization. We observed a spectrum of risk which we termed the aortic biomechanical vulnerability index (AortoBVI), onto which aneurysm patients could be mapped. This first iteration demonstrated strong discriminative performance (AUC = 0.901, 95% bootstrap CI 0.836–0.954; **Figure 3a,b**) and good calibration (Brier score = 0.129; **Figure 3d**), indicating that aortic biomechanics can reliably stratify biomechanical risk phenotypes beyond a proof of concept. AortoBVI itself may serve as the elusive aortic endpoint for future studies exploring novel biomarkers and therapeutics.

The histological associations we observed provide orthogonal biological support for this interpretation. Elastin area fraction was inversely associated with tissue-derived AortoBVI, whereas collagen area fraction was positively associated with AortoBVI (**Figure S3b,c**). Elastin loss and collagen accumulation are consistent with the extracellular-matrix remodeling in medial degeneration, supporting AortoBVI’s biological relevance beyond the mechanical measurements from which it was derived.

Of the 353 aneurysms undergoing elective surgery, 165 were categorized as dissection-like, 93 normal-like, and 95 as intermediate (**Figure 3e**). Thus, while many aneurysms were appropriately operated on, we argue that many were not. There are, therefore, patients who have suffered surgical complications for whom durations of complication-free survival were unnecessarily shortened. Aortic diameter explained only a small proportion of the variance in tissue-derived AortoBVI across the full cohort (R² = 0.075, n = 350; **Figure S8a**) and diameter strata showed 10 of 29 patients (34%) with diameter <45 mm were dissection-like, while 20 of 113 patients (17.7%) at or above the 55 mm surgical threshold remained normal-like (**Figure S8b**). These findings align with the prior clinical and translational studies describing the poor performance of maximal aortic diameter in predicting aortic risk^6,12,13^. Yet, diameter thresholds for surgical decision-making driven by isolated population-based studies currently inform all cardiothoracic surgical society guidelines^4,5^. Further shifting of size thresholds, as occurred in recent guideline iterations, is unlikely to resolve these issues^4,5^. In our cohort, at the current 55 mm diameter threshold, 93 of 165 (56.4%) aneurysms with a dissection-like AortoBVI had diameters below this cutoff. Lowering the threshold to 50 mm would still leave 37 of 165 (22.4%) dissection-like aneurysms below the threshold while increasing the proportion of normal-like aneurysms exceeding the threshold from 22.0% to 58.2% (**Figure S8c**). Here, we proposed a personalized, biomechanics-based approach to aneurysm risk assessment.

To make this risk assessment clinically tractable, we introduced a novel multi-parameter MRI-based solution that outperforms diameter in estimating AortoBVI. This MRI-AortoBVI could function as a virtual biopsy after an aneurysm diagnosis is made, providing insight into the biomechanical phenotype of the patient, thereby aiding clinical decision-making such as reducing surveillance imaging burden in low-risk patients and enabling earlier life-saving intervention in high-risk patients.

As a first step in its development, we interrogated individual MRI-based metrics for measuring aortic biomechanics in vivo. We found piecewise linear relationships with inflection points between strain, compliance and distensibility, and energy loss (EL) and delamination strength (S_d_). The inflection points may potentially be useful for dichotomizing these parameters into high- and low-risk categories. The relationships between the in vivo and ex vivo parameters were surprising and multi-pronged. For instance, strain and strain-based variables only moderately correlated with low strain modulus, despite an expected strong relationship based on their definition. This reflects the complex nonlinear stress-strain relationship of aortic tissue and the unpredictability of where each individual patient is positioned on the stress-strain curve in vivo. Notably, strain, compliance, distensibility, and ASI also correlated with EL and S_d_, two clinically relevant parameters that have been correlated with the underlying tissue microstructure and with different at-risk patient groups^13,15,16,18,35^, but on the surface are unrelated to strain. Energy loss reflects the viscoelastic properties of the ascending aorta, partly attributed to proteoglycan content^18,30,36,37^. Increased proteoglycan content is a prototypical feature of medial degeneration^36^, in which pools of excess proteoglycans generate localized Donnan swelling pressures that lead to aortic tissue delamination or dissection^38,39^. This may explain why energy loss tracks with delamination strength. Additionally, our group and others have demonstrated the interdependence of extracellular matrix components within the aortic wall, whereby proteoglycan content was found to directly influence the elastin and collagen networks by pre-straining fibers^30,37^. This interaction was hypothesized to modulate fiber recruitment and has been experimentally shown to modulate the nonlinear tensile behaviour of the tissue^30,37^. Because aortic viscoelastic and failure behaviour are impractical to directly measure in vivo, these results encouragingly suggest that the interdependency of extracellular matrix components may be leveraged and simply-obtained in vivo biomechanical properties may still capture the desired ex vivo parameters.

In contrast to the strain-based parameters, aPWV did not show a significant univariate association with ex vivo tissue mechanics in this cohort (**Figure 5m–o, Figure S6i–j**). This was initially surprising given that aPWV has well-recognized associations with atherosclerosis, hypertension, and aging^40,41^ and that previous work using carotid-femoral pulse wave velocity (cfPWV) in patients with ATAAs has demonstrated correlations with aortic growth rate^26,27,32^. One possible explanation is that aPWV was measured from the sinotubular junction to the distal descending aorta and does not reflect the ascending aorta alone. However, its ongoing utility in imaging-based biomechanical phenotyping will require iterative re-examinations.

While these associations ground the in vivo measurements in established ex vivo aortic mechanics, the objective was not to estimate any single tissue property in isolation, but to determine whether clinically accessible variables could reproduce each patient’s position along the tissue-derived vulnerability continuum (AortoBVI). The combination of age, ASI and aPWV provided the strongest approximation of tissue-derived AortoBVI (*R*² = 0.580; *P* < 0.001), outperforming all single-variable models, including maximum aortic diameter (*R*² = 0.116). The ability to non-invasively identify a dissection-like biomechanical phenotype with an AUC of 0.928 at this early stage is a convincing signal that this workflow can generate a clinically useful tool for aortic aneurysm risk stratification.

Importantly, the components of the multivariable model are captured in a single scan and require no extensive computational modeling, giving it tremendous scalability potential. MRI remains the predominant method of following young patients with thoracic aortic aneurysms who require lifelong surveillance imaging. Therefore, adoption of MRI-based aortic biomechanics lies within current clinical workflows.

Some limitations of our approach should be acknowledged. Although the ex vivo risk classifier was internally validated using nested cross-validation and bootstrap confidence intervals, its structure, thresholds, and performance estimates were all derived within a single-center dataset and therefore still require external validation in independent cohorts. Moreover, AortoBVI should be interpreted as a continuous measure of similarity along a donor-to-dissection biomechanical phenotype rather than as an absolute probability of future dissection. The extent to which this biomechanical axis determines prospective clinical events remains to be established. AortoBVI also describes the material properties of the aorta alone and should be interpreted in the context of the forces applied to that aorta. For example, one dissected aorta in our study demonstrated low AortoBVI but belonged to a patient with a resting blood pressure above 200 mmHg. Additionally, the categorical AortoBVI zones depend on the selected operating characteristics. The primary rule-out and rule-in boundaries were chosen to prioritize high sensitivity and specificity, respectively, and alternative clinically plausible boundary definitions altered the proportions of aneurysms assigned to the three zones (**Figure S2**). To address the patients who lie close to these boundaries, the predicted AortoBVI is provided as an interval rather than a singular number, therefore avoiding absolute risk zone assignment for patients for whom there is uncertainty. In relating ex vivo and in vivo parameters, aPWV was pre-selected in the multivariable model given its alternative mechanism to the stiffness-based group of features. Its incremental contribution to the multivariable imaging model (**Figure 6**), may reflect complementary, subtle information about ascending aortic biomechanics beyond stiffness-based parameters alone. However, as aPWV was not associated with tissue-derived AortoBVI in this cohort in univariate analysis, this interpretation requires caution; a larger paired cohort is needed to confirm whether aPWV meaningfully improves risk estimation beyond age and ASI. Similarly, because the paired in vivo–ex vivo cohort is small (n = 38), overfitting may compromise the regression equations. We mitigated this in two ways: first, by restricting the candidate model space to simple, prespecified, and interpretable functional forms rather than a data-driven search; and second, by reporting performance via leave-one-out cross-validation rather than in-sample fit (**Table S2, Figure 6**). Even so, the current regression coefficients and prediction intervals should be viewed as a demonstration of signal and biological plausibility.

In conclusion, we present AortoBVI, a tissue-derived, machine-learning-defined continuum of aortic biomechanical vulnerability that can be estimated non-invasively from MRI. By moving beyond diameter as an imperfect surrogate of aortic wall behavior, this framework provides a path toward personalized aneurysm assessment by identifying biomechanical phenotypes that are not captured by size alone. Because AortoBVI is built from routinely available clinical variables and a quantitative imaging workflow that could be adapted to existing surveillance imaging platforms, it offers a practical foundation for prospective validation and the future transition from population-based size thresholds toward individualized, biomechanics-informed decision-making.

## Data Availability

Data and code can be made available to reviewers, and will be made available publicly upon publication, subject to privacy and intellectual property restrictions.

## Reproducibility

The frozen deployment equation is implemented as dependency-free arithmetic requiring only NumPy and is automatically verified against the fitted pipeline at each build.

## Author Contributions

J.C.Y.C. and C.A.S. conceived the study. J.C.Y.C. and M.O. performed the surgical procedures and contributed clinical expertise and patient specimens. D.E. and H.Seo performed the biomechanical and histological experiments. F.T., N.B., D.H and H.Su processed the MRI data. F.T. developed the machine-learning models and performed the statistical and machine-learning analyses, with methodological input from C.M. J.C.Y.C., F.T. and D.E. contributed to data curation. K.H. provided radiological expertise, imaging resources and support for the MRI analyses. C.A.S. provided resources for biomechanical and histological testing. J.C.Y.C. and C.A.S. supervised the study. J.C.Y.C. and F.T. wrote the original draft. All authors reviewed and edited the manuscript. J.C.Y.C. and C.A.S. acquired funding for the study.

## Acknowledgements

This work was supported by Canadian Institutes of Health Research (CIHR), the American Association for Thoracic Surgery Surgical Investigator Program, the University of Toronto EMHSeed Award, the Canada First Research Excellence Fund through the Medicine by Design initiative, and the Melanie Munk Professorship in Aortic Biomechanical Research.

## Conflicts of interest

The authors have no relevant conflicts.

## Sources of funding

Canadian Institutes for Health Research (CIHR), American Association for Thoracic Surgery (AATS) Surgical Investigator Program, University of Toronto EMHSeed Award, the Canada First Research Excellence Fund (CFREF) Medicine by Design initiative, Melanie Munk Professorship in Aortic Biomechanical Research,

## Notes

### Competing Interest Statement

The authors have declared no competing interest.

### Author Declarations

This prospective cohort study was approved by the research ethics board of the University Health Network, Toronto (16-6285, July 27, 2017).

